# Evaluation of Trans-Sodium Crocetinate (TSC) on Peripheral Oxygenation in Healthy Individuals Using Transcutaneous Oximetry

**DOI:** 10.1101/2022.01.16.22269147

**Authors:** Martin Kankam, Stacy Handley, Chris Galloway, Dick Clarke

**Affiliations:** Co-Medical Director, Altasciences Clinical Kansas Inc., Overland Park, KS; VP National Baromedical Services, Columbia, SC; Chief Medical Officer, Diffusion Pharmaceuticals Inc, Charlottesville, VA; President, National Board of Diving & Hyperbaric Medical Technology, Columbia, SC

## Abstract

**Introduction:** Trans sodium crocetinate (TSC) is a synthetic carotenoid with a unique mechanism of action that improves the diffusion of oxygen by reducing oxygen transfer resistance within plasma, and it is currently being developed to enhance oxygen delivery to hypoxic tissues in multiple conditions. The goals of this study were to evaluate safety, pharmacokinetics, and pharmacodynamic properties of escalating doses of TSC on peripheral oxygenation utilizing transcutaneous oxygen measurements, when administered to healthy subjects breathing supplemental oxygen.

**Methods:** This was a dose-escalation, single-center, randomized, phase 1 study aimed at assessing the safety, pharmacokinetic and pharmacodynamic properties of TSC at doses of 0.5, 1.0, 1.5, 2.0, or 2.5 mg/kg as an intravenous bolus. Thirty healthy adult subjects of 18 to 55 years of age were enrolled and allocated to one of the five dose groups or placebo. Venous blood samples were collected for pharmacokinetic evaluations of TSC at 1, 10, 30 minutes, and 1.5 hours after the start of injection of the study drug. Pharmacodynamic assessment of tissue oxygenation was performed while the subjects breathed supplemental oxygen at 6 L/minute for 70 minutes prior to study drug administration: the first 10 minutes was to allow for equilibration, and the subsequent 60 minutes served as a baseline period (Period 1), followed by a time-matched 60-minute intervention period (Period 2). Tissue oxygenation readings were obtained by transcutaneous oximetry (TcpO_2_) measurement using four TcpO_2_ sensors placed on the lower limbs of subjects lying in a supine or semi-recumbent position. TcpO_2_ values were recorded over a 2-hour time period: 60 minutes prior to study drug administration (Period 1) and 60 minutes post administration of the study drug (Period 2).

**Results:** TSC was safe and well tolerated at all doses tested. The pharmacokinetic analyses demonstrated that clearance decreased at escalating doses of TSC. The results of the primary pharmacodynamic analysis revealed high levels of variability in the 60-minute baseline TcpO_2_ levels, however despite such variability, time-matched TcpO_2_ measurements demonstrated observed increases in median TcpO_2_ values in subjects who received TSC, relative to those who received a placebo. The high variability observed across the four sensors suggested that the data could not be pooled across all four sensors, therefore, additional supplemental analyses were performed. The results of the supplemental analyses indicated that the TcpO_2_ intra-subject slopes of the TSC treatment groups were consistently positive during the study intervention period, and therefore suggestive of an increase in TcpO_2_ levels. This was not observed in the placebo group. Based on this analysis, all TSC dose groups had a greater increase in TcpO_2_ levels than the placebo group, with the 2.5 mg/kg dose demonstrating the most notable increase over the 1-hour intervention period (Period 2).

**Conclusions:** TSC administered as a single IV bolus dose ranging from 0.5 mg/kg to 2.5 mg/kg to healthy subjects breathing supplemental oxygen, was safe and well tolerated. Pharmacokinetic assessments demonstrated that TSC plasma concentrations increased with escalating dose and that increasing TSC dose was associated with a decrease in clearance. The high levels of variability in TcpO_2_ levels did not allow for pooling of sensor measurements for primary analysis; however, supplemental analysis of individual sensor measurements demonstrated an observed dose effect of TSC on peripheral tissue oxygenation relative to placebo.

## Introduction

Oxygen is essential in mammals for aerobic metabolism and therefore the proper delivery of oxygen to tissues is vital (1). Delivery of oxygen to tissues is dependent on both convection and diffusion. Convective delivery of oxygen is primarily driven by cardiac output and arterial oxygen content, while the diffusive transport occurs at the microcirculatory level and is dependent on Fick’s Law of diffusion, therefore representing a rate limiting step (2). Gainer *et al*. demonstrated that crocetin, a natural carotenoid compound, was able to increase the diffusion of oxygen through plasma (3,4).

When local, regional, or generalized oxygen delivery to tissues is either inadequate or disrupted, cells become hypoxic; if not reversed, hypoxia can rapidly lead to tissue damage and cell death (1). Stroke, myocardial infarction, COVID-19 are just some examples of conditions complicated by hypoxia, for which hypoxia directly contributes to short- and long-term morbidity and mortality. In addition, hypoxia can often complicate many sub-acute conditions, such as with many cancers (6). Hypoxia is present in nearly all solid tumors and has been shown to contribute to resistance to radiotherapy, chemotherapy, and immunotherapy, as well as increase the potential for tumor invasion and metastasis (5).

Diffusion Pharmaceuticals has developed trans sodium crocetinate (TSC), a fully synthetic *trans*-isomer salt of crocetin that has demonstrated the ability to increase the diffusion of oxygen by transiently increasing hydrogen bonds between water molecules within the plasma component of blood (4). This enhances the organizational matrix within plasma, which facilitates the passive diffusion of oxygen from high to low concentration areas with less resistance (6,7).

*In-vitro* studies have shown that TSC increases oxygen diffusion in water by up to 30% (8,9). Following these *in-vitro* experiments, *in-vivo* studies were performed to assess the potential of TSC as a therapeutic intervention in animal models of hypoxia. Several studies were performed in various ischemic animal models where TSC was administered following ischemic shock, and these studies demonstrated significant improvement in survival rates of rats recovering from post ischemic shock (12–15). TSC was also shown to enhance radiosensitivity in a glioblastoma multiforme (GBM) model in rats (16), and to improve survival in a rat model of hypoxemia (17).

The safety and tolerability of TSC was first clinically evaluated in a placebo-controlled Phase 1 study enrolling 40 normal healthy subjects (15). The results of this study demonstrated that TSC was safe and well tolerated with a maximum tolerated dose of 2.5 mg/kg. Safety, tolerability, and dose response were also evaluated in a placebo-controlled Phase 1/2 study of 48 patients with symptomatic peripheral artery disease (PAD) with claudication (16,17). Multiple doses of TSC were demonstrated to be safe and well tolerated, with improved scores observed at higher doses in a test of peak walking time and in-patient perceived walking distance. TSC has been evaluated for safety and dose tolerance in a placebo-controlled Phase 1/2 study of 59 patients with newly diagnosed GBM who also received chemotherapy and radiation therapy standard of care (18). The results of this study also demonstrated that TSC was safe and well tolerated. In 2020-2021, TSC was evaluated in a Phase 1/2 study of 24 patients with confirmed SARS-CoV-2 infection and hypoxemia who required hospitalization. The results of this study demonstrated that administration of TSC every six hours for up to 15 days was safe and well tolerated with clinical benefit observed on the WHO Ordinal Scale for Clinical Improvement and length of stay in the treatment of COVID-19-related hypoxemia at the highest doses tested in the trial (19).

Along with safety, the objectives of this study were to evaluate pharmacokinetic and pharmacodynamic responses after administration of TSC or placebo in a randomized and blinded trial in the peripheral tissues of healthy subjects breathing supplemental oxygen, as measured by transcutaneous oximetry (TcpO_2_). Transcutaneous oximetry is a non-invasive test which measures the partial pressure of oxygen diffusing through the skin and provides insight into local tissue oxygenation (20). The intent was to further characterize the objective dose-response relationship of TSC and tissue oxygenation to provide data for further studies.

## Methods

### Design and Dosing

This was a single center randomized, double-blind, placebo-controlled, pharmacokinetic and pharmacodynamic study of TSC in 30 subjects. Healthy, non-smoking male and non-pregnant female volunteers aged 18 to 55 years with a body mass index ranging from 18.0 to 30.0 kg/m^2^ and a minimum body weight of 50.0 kg were eligible for study inclusion. A screening visit occurred within 21 days of drug administration. Health status was determined by medical history, physical examination, oxygen saturation, electrocardiogram, laboratory tests and urine drug screen. A negative serum pregnancy test was required for women. Subjects were instructed to refrain from exercise, caffeine, alcohol, and a heavy meal prior to study drug administration. On the treatment day, the inclusion and exclusion criteria were reassessed and a pregnancy test, drug screen, and alcohol screen were performed. Mean (±SD) age was 33 (±9) years (Table 1). The subject population included non-smokers, different ethnicities, and a balanced ratio of males and females. One subject in the 2.5 mg/kg dose cohort experienced inadvertent subcutaneous infiltration of TSC due to IV cannula migration and was excluded from the analysis. Study results reflect the pharmacokinetic and pharmacodynamic analyses of 24 and 29 subjects, respectively.

**Table 1:**
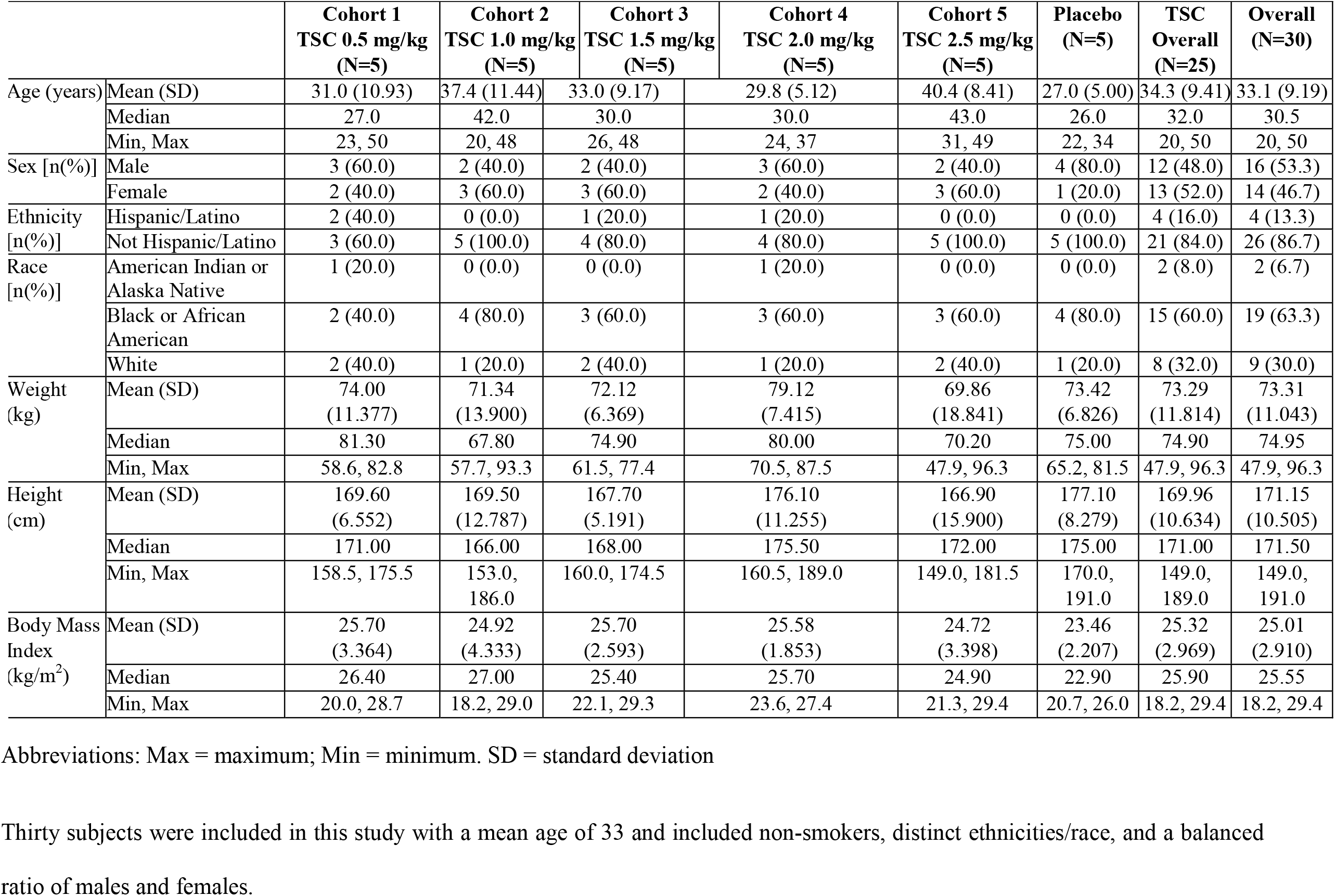
Subject Demographics.

The study was conducted in a temperature-controlled room (22.0-25.0°C). Subjects maintained a supine or semi-recumbent position and were asked to lie quietly and minimize body movement for the entire study procedure.

Tissue oxygenation was assessed by applying four transcutaneous oximetry sensors to a lower limb of each subject. Sensors were connected to a transcutaneous monitor (Radiometer America, Inc., Brea CA 92821) set to 45° C. Sensor locations were: mid-dorsum of the foot (Sensor 1); 10 cm distal to the lateral femoral epicondyle (Sensor 2); 5 cm proximal to the anterior aspect of the lateral malleolus (Sensor 3); and 5 cm proximal to the anterior of the medial malleolus (Sensor 4).

Subjects breathed supplemental oxygen (6 L/min, administered via simple facemask) beginning 10 minutes before recording of TcpO_2_ measurements for equilibration, and then the next 60 minutes comprised the baseline period (Period 1). At the end of Period 1, subjects received an IV bolus injection of TSC or placebo as a single intravenous dose at time zero; and then continued with a 60-minute treatment observation period (Period 2). TcpO_2_ and SpO_2_ values were recorded every five minutes during Period 1. Each cohort included five subjects administered TSC (0.5, 1.0. 1.5, 2.0, or 2.5 mg/kg) or placebo (7 ml normal saline).

Period 2 transcutaneous oximetry readings included observations at 1-, 3-, and 5-minute intervals after TSC dosing, then every 5 minutes thereafter. Vital signs (heart rate, blood pressure, respiratory rate) were measured at clinic arrival, baseline, and at 10, 30, and 60-minutes post study drug dosing. Subjects were monitored for adverse events for the study duration, remaining in the research unit overnight, and were contacted by telephone 48 hours after injection of study drug or placebo to assess adverse events and any new medication use.

### Pharmacokinetic Sampling and Analysis

Venous blood samples were drawn within 10 minutes before study drug injection and at 1, 10, 30 minutes (±1 minute), and 90 (± 2) minutes after the injection to determine the concentration of TSC.

Plasma was assayed for TSC using a validated liquid-chromatography-tandem mass spectrometry (LC-MS/MS) analytical method. Sample pre-treatment involved a protein precipitation extraction procedure. The LC-MS/MS method utilized electrospray ionization in the negative ionization mode, with prednisone as an internal standard. The lower limit of quantitation (LLOQ) was 10 ng/mL.

A population pharmacokinetic analysis was performed using the nonlinear mixed-effects modeling program NONMEM (version 7.4.3, ICON Development Solution, Ellicott City, MD). Based on previous studies with TSC and examination of the concentration profiles, a two-compartment linear model was selected. The weight normalized model was preferred statistically, after consideration of other covariates’ effect on the model (i.e., age, sex, organ impairment, and dose level).

### Pharmacodynamic Analysis of Peripheral Tissue Oxygenation

The overall pooled median values across the four TcpO_2_ sensors were calculated using the median value from each sensor during the same time interval. To compare the time-matched changes in TcpO_2_, a 2-factor (treatment and time) repeated measures (time) analysis of variance (ANOVA) model was used. Contrast statements within the model facilitated comparisons between individual dose arms and placebo over time. Intra-subject changes (Period 2 minus Period 1 in 5-minute intervals) served as the dependent variable in the model. TcpO_2_ baseline measures during the 60-minute Period 1 adjusted for intra-subject variability over time. Time-matched differences between values obtained during Period 1 and Period 2 represented the least biased estimate of the effect of the study drug versus placebo.

### Supplemental Analysis of Tissue Oxygenation Pharmacodynamics

The raw data recordings of the individual TcpO_2_ sensor measurements demonstrated high variability among sensors, suggesting that the data should not be pooled across all four sensors. Therefore, a supplementary pharmacodynamic analysis on the intra-subject slopes from each period and sensor was performed. For Period 1, on an intra-subject basis, the median value from each of the 12 intervals was used to construct the slope using orthogonal spacing with respect to time. For Period 2, the slope was constructed using the median value from each of the 13 intervals—of note, Period 2 observations included 1- and 2-minute values—in addition to the 5-minute measurements for comparison to the 12 corresponding intervals of Period 1. An additional set of analyses were conducted using the Period 2 intra-subject slopes as the dependent variable. The active dose groups were pooled and the least square means were compared to placebo. This procedure was repeated iteratively, each time removing the next lowest dose group to determine the separation of each cohort’s least square means relative to placebo.

## Results

### Pharmacokinetic Analysis

For 19 of the 24 subjects included in the analysis, TSC concentration in the 1-minute sample was smaller (often many-fold) compared to the 10-minute sample (Figure 1), most likely a result of sampling too soon after the dose was administered. As a result, all 1-minute samples were excluded from the NONMEM population Pharmacokinetic analysis.

**Figure 1:**
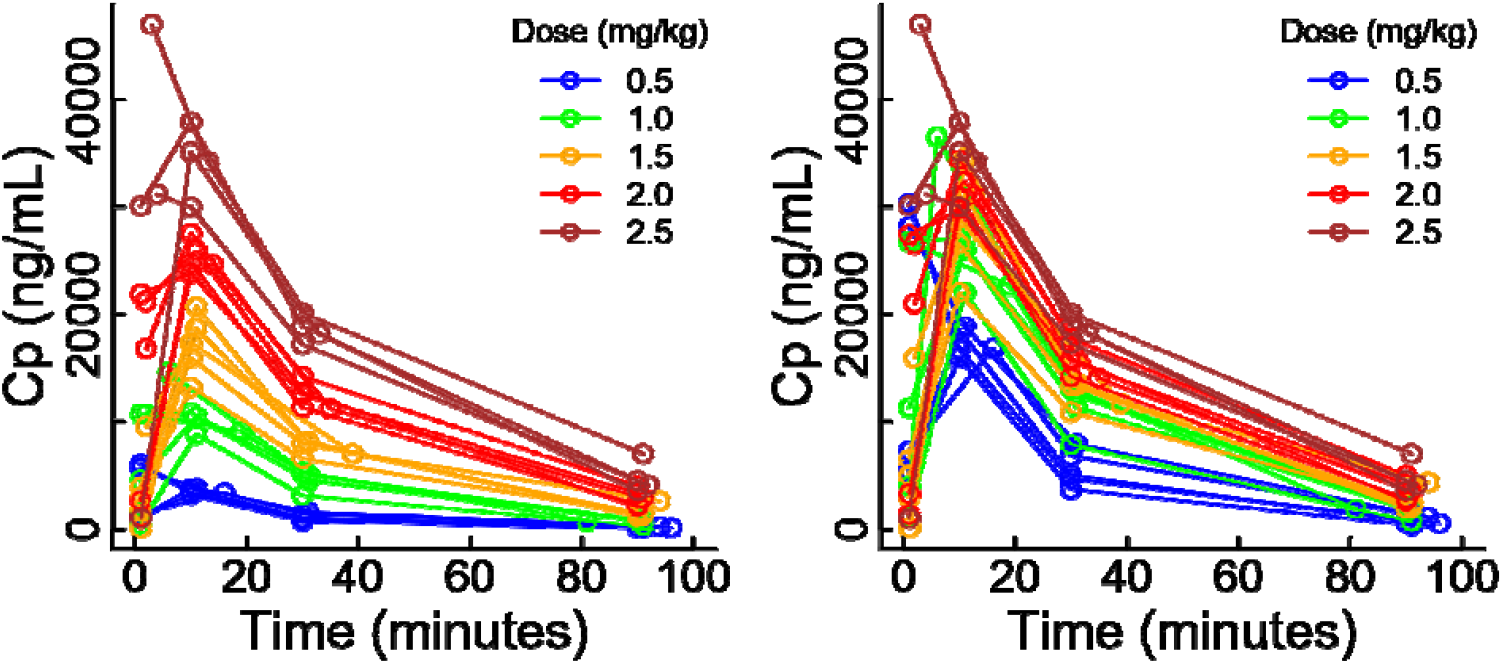
TSC Concentration Profiles for Each Subject. TSC plasma concentrations increase with escalating dose, with the 2.5mg/kg dose administration leading to the highest plasma concentration. Raw values are presented in the left panel and concentrations normalized to a TSC dose of 2.5 mg/kg are presented in the right panel. Cp: Concentration in Plasma. TSC concentration in the 1-minute sample was smaller than the 10-minute sample, most likely a result of sampling too soon after the dose was administered.

Based on previous analyses and examination of drug concentration profiles, a two-compartment model with no covariates fit the data well. To determine whether the systemic parameters should be scaled for body size, three scaling approaches were evaluated: scaling all parameters by weight, allometric scaling (clearances scaled by weight raised to the 0.75 power; distribution volumes scaled by weight), and scaling all parameters by weight raised to an estimated power. The weight normalized model was preferred statistically (evaluated by the decrease in the objective function [similar to a sum of squares]) and by graphics. Graphics suggested that clearance varied as a function of dose. A model was evaluated in which clearance varied as a linear function of dose:

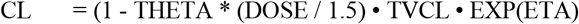

where CL, each subject’s individual (*post hoc*) value for clearance, is the product of linear function of dose (where DOSE is the dose in mg/kg and THETA is estimated), 1.5 is the median dose (thereby centering the model), TVCL is the “typical” value for clearance (which applies to subjects receiving the 1.5 mg/kg dose, and EXP(ETA) is a term that allows for inter-individual variability. Pharmacokinetic parameters are displayed in Table 2. The concentration profile for a “typical” subject (weighing 75 kg) administered escalating drug doses from 0.5 - 2.5 mg/kg is displayed in Figure 2.

**Table 2:** Population Pharmacokinetic |Parameters of TSC.

| Description | Estimate | Inter-individual Variability* |
| --- | --- | --- |
| SCALE | (WT/75)<sup>†</sup> |  |
| FCTR | $1 - 0.53677 \cdot (\text{DOSE}^{\S} / 1.5)$ | —<sup>¶</sup> |
| Clearance (L / day) | $196.561 \cdot \text{SCALE} \cdot \text{FCTR}$ | 0.6189 |
| V<sub>1</sub> (L) | $4.27707 \cdot \text{SCALE}$ | 0.1869 |
| Distribution Clearance (L / day) | $161.176 \cdot \text{SCALE}$ | 0.1296 |
| V<sub>2</sub> (L) | $46.61 \cdot \text{SCALE}$ | 0.6743 |

\* Quantified as $\sqrt{\text{OMEGA}^2}$ where $\text{OMEGA}^2$ is the variance of inter-individual variability.
<sup>†</sup> WT is weight in kg; 75 kg was the median weight in the study
<sup>§</sup> DOSE is dose in mg/kg; 1.5 is the median value
<sup>¶</sup> Inter-individual variability was not permitted for this term.

**Figure 2:**
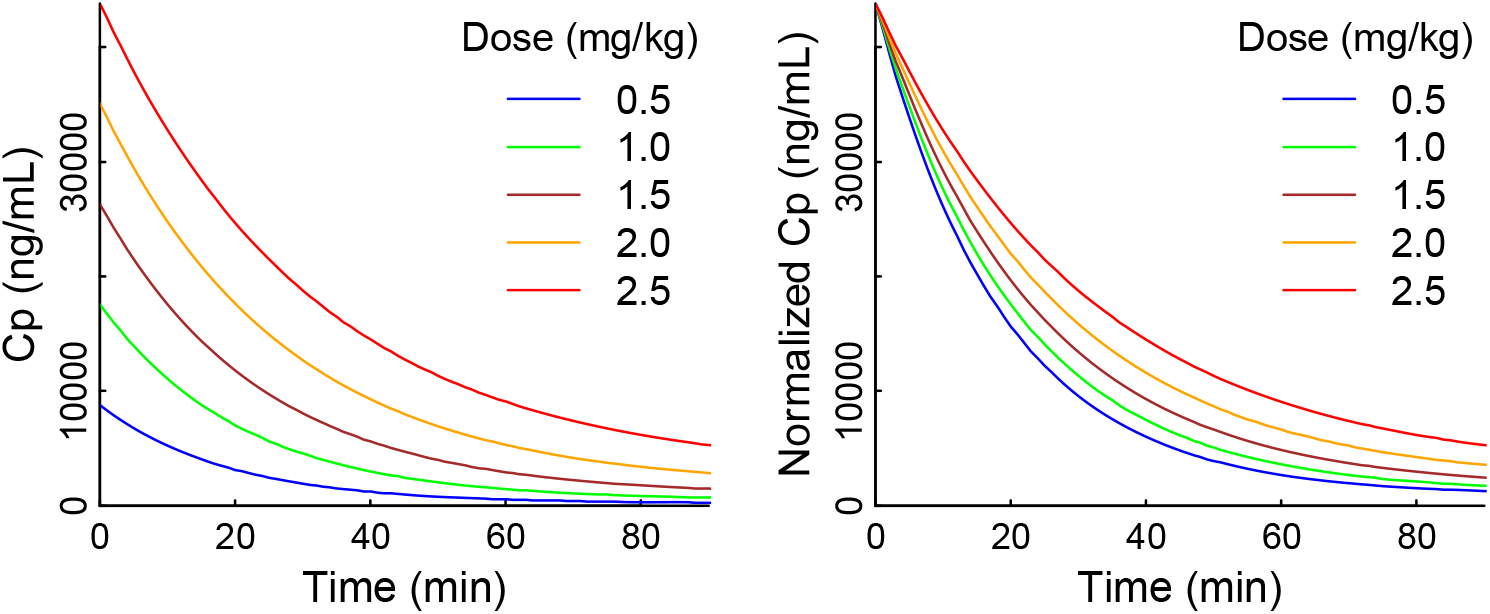
Simulated TSC concentration profiles. Simulated TSC concentration profiles for a subject weighing 75 kg, based on the population parameters in Table 2. In the right panel, concentrations are normalized to a dose of 2.5 mg/kg to illustrate the non-linearity with respect to dose.

### Pharmacodynamic Analysis

Baseline TcpO_2_ values recorded in this study were between 150-200 mmHg which is in line with previous studies of healthy individuals (21,22). The time-matched TcpO_2_ values reporting changes in the amount of oxygen in tissue extremities fluctuated during baseline Period 1, an unexpected finding. The high variability in the baseline Period 1 TcpO_2_ readings did not clearly demonstrate time matching trends; however, the placebo group displayed a time related decrease in peripheral oxygenation during Period 2. Conversely, the TSC groups displayed stable or increasing peripheral oxygenation levels over time (Figure 3). Furthermore, Period 2 assessments of the difference in tcpO_2_ values between TSC and placebo (Figure 4) suggested an increase in peripheral TcpO_2_ values following TSC treatment in the subject groups administered higher TSC doses (≥2mg/kg).

**Figure 3:**
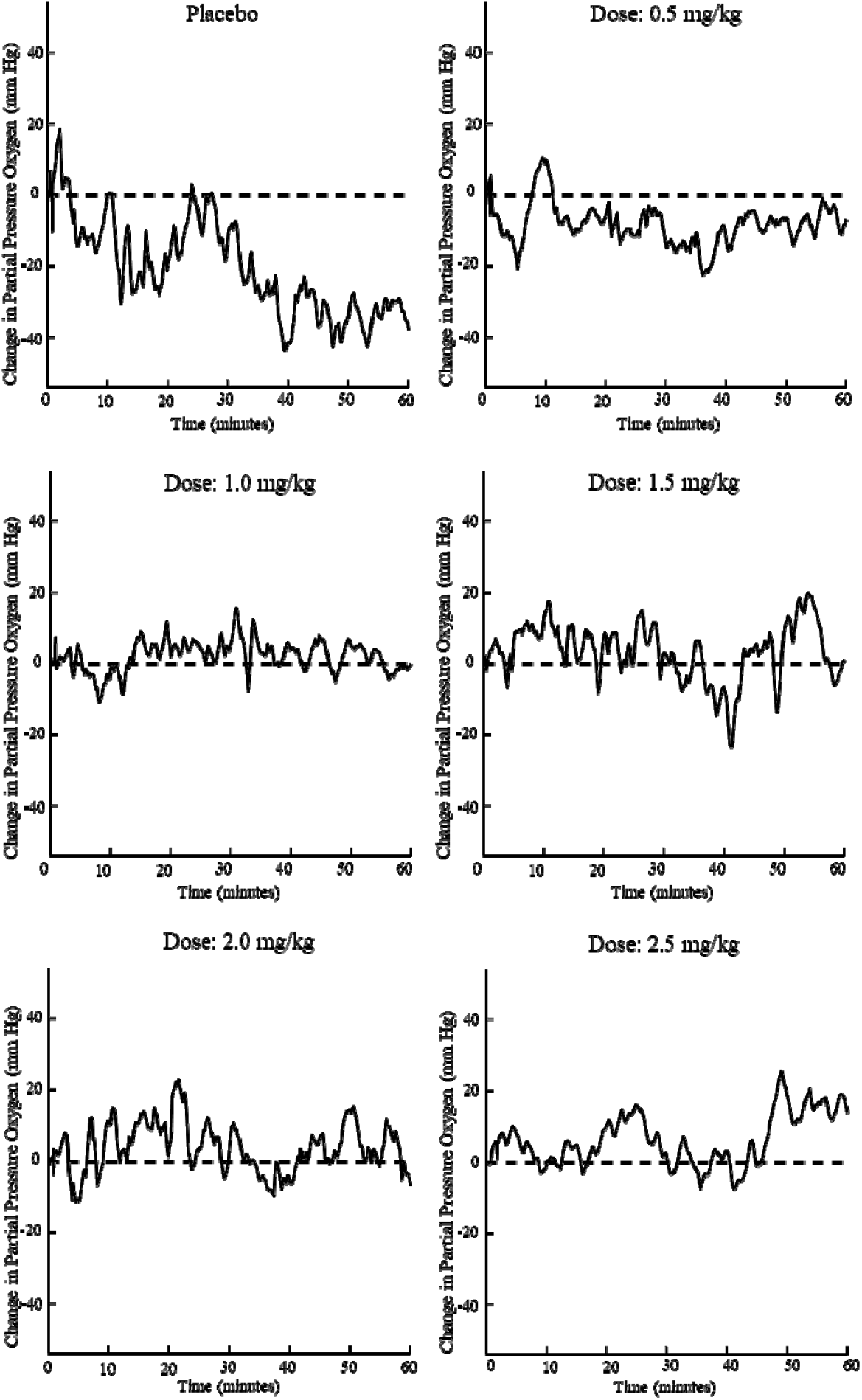
Median TcpO_2_ values during Period 2. The figure above shows changes from baseline in the median transcutaneous oxygen tension values across all dose levels. A time related decrease in peripheral oxygenation in the placebo group is observed and in contrast stable peripheral oxygenation levels over time in TSC treated groups.

**Figure 4:**
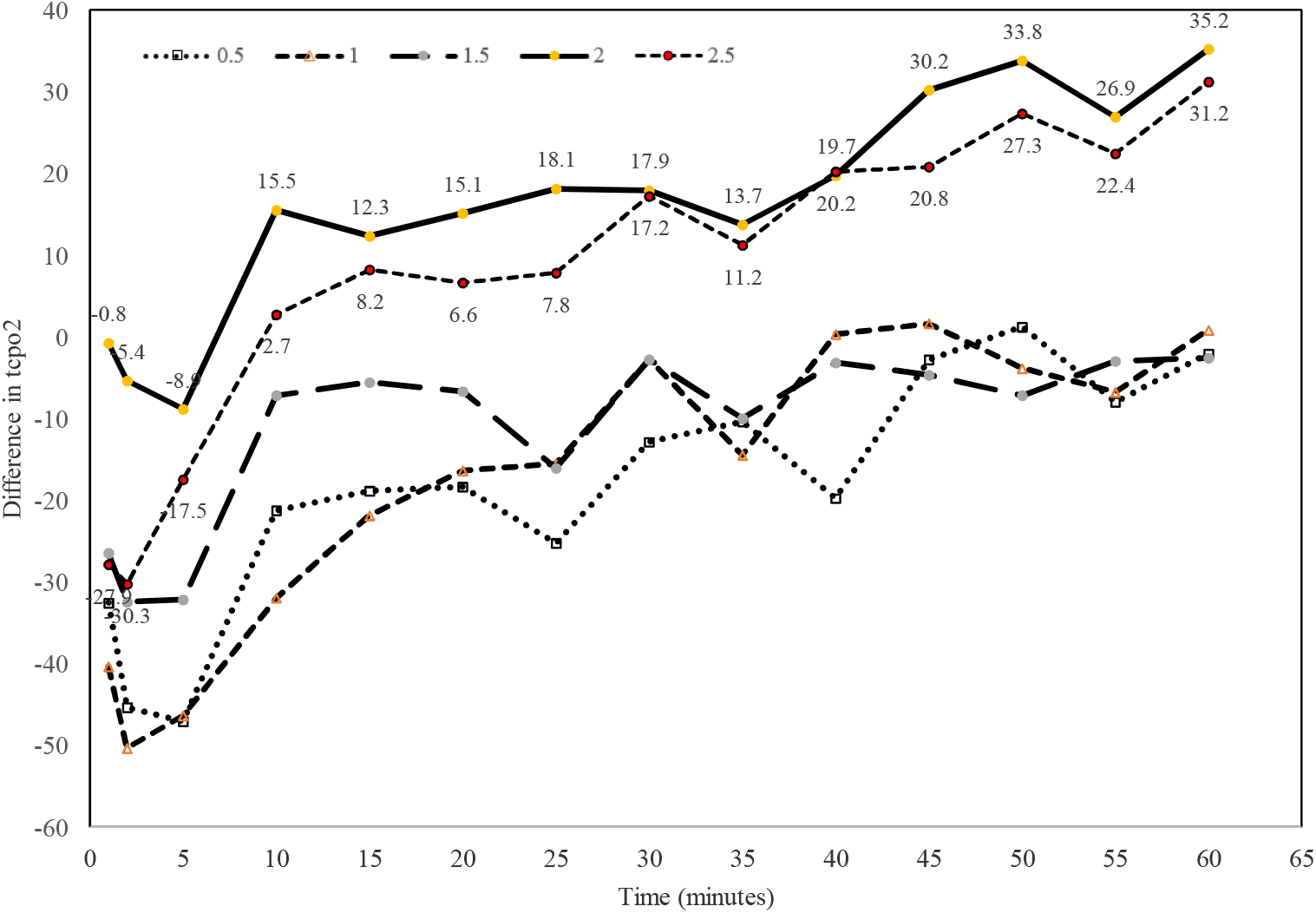
Median Change in TcpO_2_ values TSC Dose minus Placebo, Period 2. The subtraction of the placebo TcpO_2_ levels from the TSC treated groups in Period 2 illustrate an increase in TcpO_2_ values post TSC administration relative to placebo in the higher TSC dose groups (≥2mg/kg). For Period 2, the median value was used from each of the 13 intervals of 1, 3-and 5-minute duration.

The high range of variability observed in the 60-minute baseline portion (Period 1) among the four sensors suggested that the data was not able to be pooled across all four sensors. Therefore, additional supplemental analyses were performed as described in the Methods section. The supplemental analyses demonstrated that the subjects of the TSC groups had consistently positive intra-subject slopes across multiple sensors observed during Period 2 that were not observed for the placebo group (Table 3). In addition, pooling the TSC groups and comparing their least square means to those of the placebo arm within Period 2 revealed a significant separation across all four sensors (Tables 4-7). The greatest least square mean difference occurred with 2.5 mg/kg dose versus to placebo.

**Table 3:**
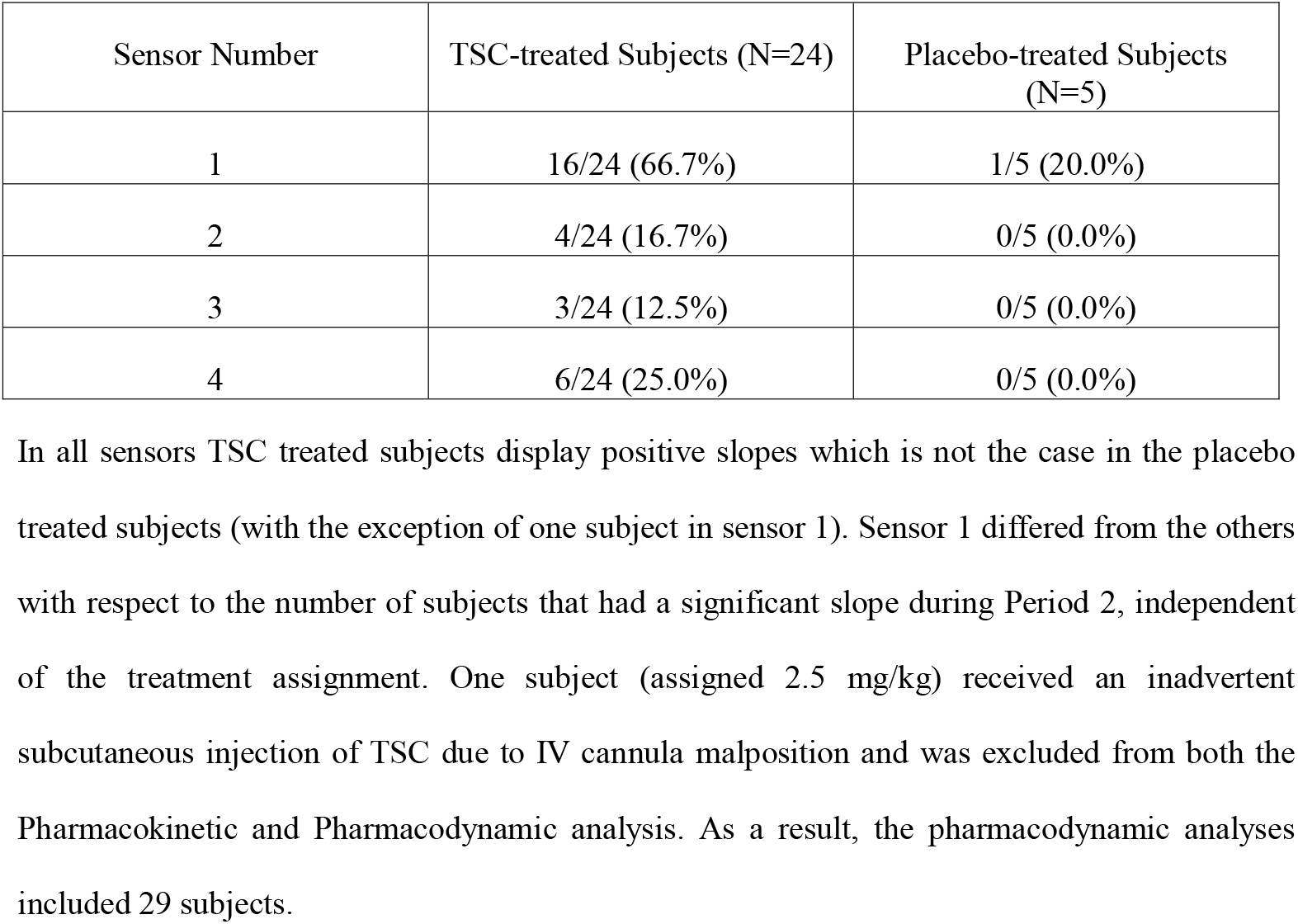
**Number (%) of Subjects with a TcpO_2_ increase during Period 2, Treatment vs. Placebo**

| Sensor Number | TSC-treated Subjects (N=24) | Placebo-treated Subjects (N=5) |
| --- | --- | --- |
| 1 | 16/24 (66.7%) | 1/5 (20.0%) |
| 2 | 4/24 (16.7%) | 0/5 (0.0%) |
| 3 | 3/24 (12.5%) | 0/5 (0.0%) |
| 4 | 6/24 (25.0%) | 0/5 (0.0%) |

In all sensors TSC treated subjects display positive slopes which is not the case in the placebo treated subjects (with the exception of one subject in sensor 1). Sensor 1 differed from the others with respect to the number of subjects that had a significant slope during Period 2, independent of the treatment assignment. One subject (assigned 2.5 mg/kg) received an inadvertent subcutaneous injection of TSC due to IV cannula malposition and was excluded from both the Pharmacokinetic and Pharmacodynamic analysis. As a result, the pharmacodynamic analyses included 29 subjects.

**Table 4:** **Summary of the Intra-Subject Slopes from Sensor 1 during Period 1 and 2 by Randomized Treatment Assignment**

| Sensor 1 | Groups being Compared | Least Square Mean | Probability Value |
| --- | --- | --- | --- |
| Pooling all TSC doses vs. placebo | TSC | 0.3185 | 0.0876 |
|  | Placebo | -0.1878 |  |
| Pooling TSC doses $\geq 1.0$ mg/kg vs. placebo | TSC | 0.3537 | 0.0960 |
|  | Placebo | -0.1878 |  |
| Pooling TSC doses $\geq 1.5$ mg/kg vs. placebo | TSC | 0.3027 | 0.1702 |
|  | Placebo | -0.1878 |  |
| Pooling TSC doses $\geq 2.0$ mg/kg vs. placebo | TSC | 0.4359 | 0.1430 |
|  | Placebo | -0.1878 |  |

Differences between the least square means of the placebo group and the TSC dose groups reveal that TSC groups have a greater trajectory than the placebo group.

**Table 5:** **Summary of the Intra-Subject Slopes from Sensor 2 during Period 1 and 2 by Randomized Treatment Assignment**

| Sensor 2 | Groups being Compared | Least Square Mean | Probability Value |
| --- | --- | --- | --- |
| Pooling all TSC doses vs. placebo | TSC | 0.0849 | 0.0796 |
|  | Placebo | -0.3481 |  |
| Pooling TSC doses $\geq 1.0$ mg/kg vs. placebo | TSC | 0.0656 | 0.1139 |
|  | Placebo | -0.3481 |  |
| Pooling TSC doses $\geq 1.5$ mg/kg vs. placebo | TSC | 0.0149 | 0.1306 |
|  | Placebo | -0.3481 |  |
| Pooling TSC doses $\geq 2.0$ mg/kg vs. placebo | TSC | 0.0753 | 0.0959 |
|  | Placebo | -0.3481 |  |

Differences between the least square means of the placebo group and the TSC dose groups reveal that TSC groups have a greater trajectory than the placebo group.

**Table 6:**
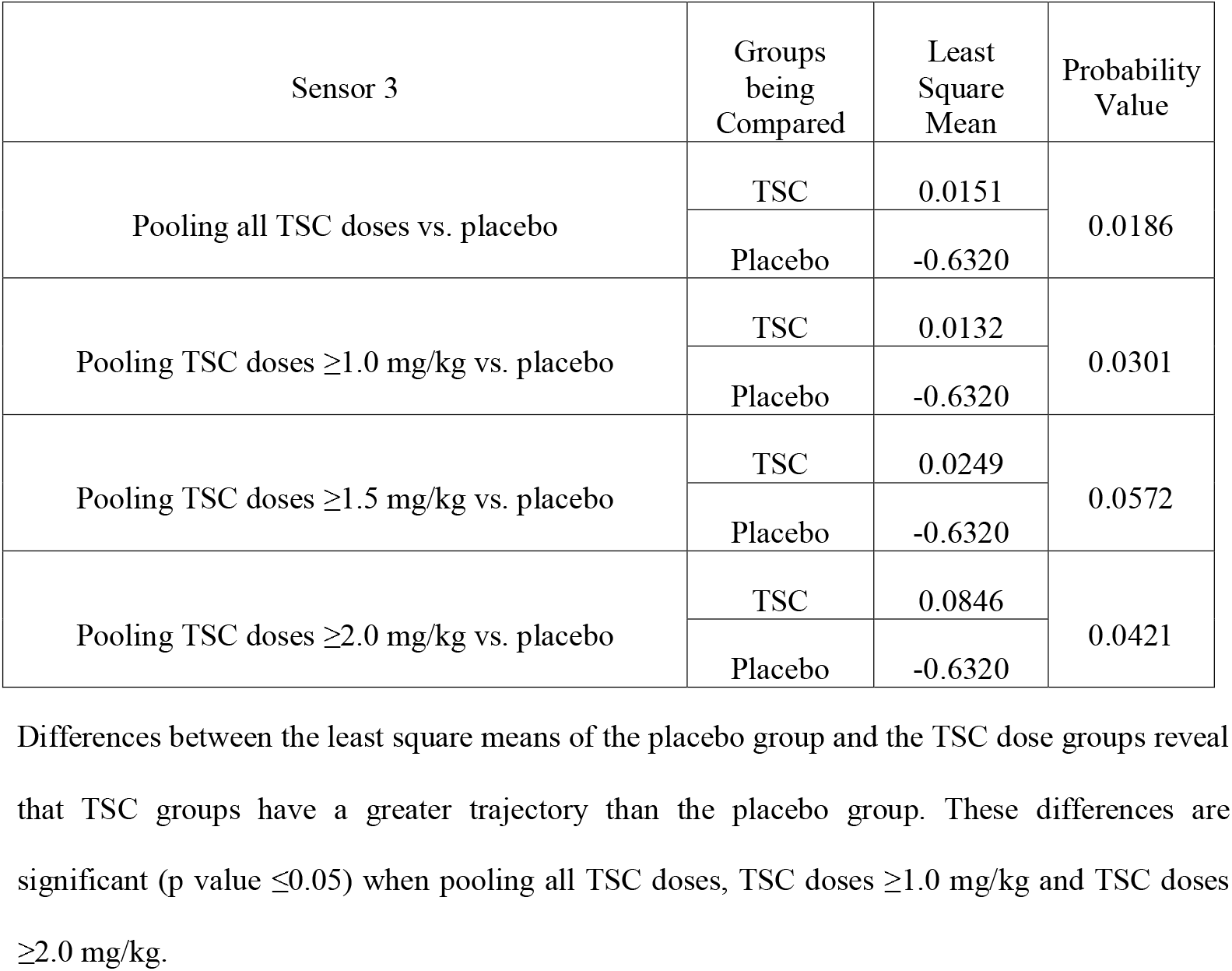
**Summary of the Intra-Subject Slopes from Sensor 3 during Period 1 and 2 by Randomized Treatment Assignment**

**Table 7:** **Summary of the Intra-Subject Slopes from Sensor 4 during Period 1 and 2 by Randomized Treatment Assignment**

| Sensor 4 | Groups being Compared | Least Square Mean | Probability Value |
| --- | --- | --- | --- |
| Pooling all TSC doses vs. placebo | TSC | -0.0198 | 0.0230 |
|  | Placebo | -0.6389 |  |
| Pooling TSC doses $\geq 1.0$ mg/kg vs. placebo | TSC | 0.0260 | 0.0213 |
|  | Placebo | -0.6389 |  |
| Pooling TSC doses $\geq 1.5$ mg/kg vs. placebo | TSC | -0.0909 | 0.0436 |
|  | Placebo | -0.6389 |  |
| Pooling TSC doses $\geq 2.0$ mg/kg vs. placebo | TSC | 0.0297 | 0.0187 |
|  | Placebo | -0.6389 |  |

Differences between the least square means of the placebo group and the TSC dose groups reveals that TSC groups have a greater trajectory than the placebo group. These differences are significant (p value $\leq 0.05$ ) in all groups.

Differences were observed in sensor 1 which had a greater number of positive intra-subject slopes and increased least square means compared to the other three sensors (Tables 3 and 4). However, the results of sensor 1 were similar to those of the other sensors, in which the TSC treated groups displayed increases in TcpO_2_ slopes which were not observed in the placebo group. Furthermore, sensor 1 was the most distal sensor in all subjects (placed on the mid-dorsum of the foot) which may in part explain the larger observed change in the TcpO_2_ levels at this sensor’s anatomic location. Augmentation in peripheral oxygenation would potentially be most notable when starting from a lower baseline level of tissue oxygenation which can occur in the most distal parts of the body, and it is conceivable that more significant increases in TcpO_2_ values would be observed in patient populations who are hypoxic at baseline.

Overall, the supplemental analysis was suggestive of an effect across all sensors in which the TSC groups had a greater increase in TcpO_2_ than placebo. Of note, the 2.5 mg/kg dose resulted in the greatest difference from placebo over the 1-hour treatment period (Period 2). There was no correction for multiplicity across the analyses, given the probability values were calculated for informational purposes.

### Safety

TSC was safe and well tolerated, with a total of nine treatment-emergent adverse events reported overall among seven of the 30 subjects (23.3%). Of the nine treatment-emergent adverse events experienced by five subjects (16.7%), five were determined to be drug-related and all were deemed mild in intensity. The most frequent treatment-emergent adverse event was post procedural erythema experienced by two subjects; others included transient chromaturia (a known effect of TSC), post procedural pruritus, headache, taste disorder, injection site pain, and injection site streaking. No subjects experienced a serious adverse event or withdrew from the study for any reason.

## Discussion

This study was designed to evaluate the dose-response effect of escalating doses of TSC on peripheral tissue oxygenation, evaluated using transcutaneous oximetry measurements in healthy subjects breathing supplemental oxygen. Overall, TSC administered as a single IV bolus dose ranging from 0.5 mg/kg to 2.5 mg/kg to healthy subjects breathing supplemental oxygen at 6 L/min was safe and well tolerated. TSC demonstrated decreasing drug clearance with increasing doses which is consistent with previous clinical studies (19).

The results of the pharmacodynamic analysis displayed a time related decrease in peripheral oxygenation during Period 2 in the placebo group which was not observed in the TSC groups. Hypotheses for this observation might include: reduction in minute ventilation and/or reduction in cardiac output in the second hour with subjects being in a semi-recumbent position and relaxing conditions. Importantly the TSC treated groups treated in identical conditions did not display such a decrease in peripheral oxygenation across all TSC doses. In addition, the results of the supplementary pharmacodynamic analysis on TcpO_2_ measurements during Period 2 revealed increases in median TcpO_2_ values in subjects who received TSC compared to placebo.

Limitations of this study include the small sample size and the observed variability across individual TcpO_2_ sensors that did not allow for pooling of sensor readings for a direct time-matched comparison. Therefore, a sensor-by-sensor supplemental pharmacodynamic analysis was performed, first calculating the slope of median values of intra-subject TcpO_2_ interval measures across individual sensors during Period 1, then constructing intra-subject slopes using the TcpO_2_ interval measures of Period 2. Additional analysis of the Period 2 intra-subject, individual sensor slopes to the least square means of the active dose cohorts-versus-placebo calculations demonstrated consistent positive increases in TcpO_2_ readings across multiple individual sensors in Period 2 for all TSC groups when compared to the placebo group. The greatest TcpO_2_ difference was observed when the 2.5 mg/kg TSC group was compared to the placebo group, suggesting that the effect of TSC is greatest at higher doses. The safety profile of TSC in this study is consistent with previous clinical experience as well as the pharmacokinetic profile between clearance and intravenous dosing of TSC.

In conclusion, TSC is a safe and novel oxygenation enhancing compound that has the potential to be a valuable adjunct to standards of care across a wide range of acute and chronic conditions complicated by hypoxia, agnostic of causation, where improving oxygen delivery is pivotal to improving patient outcomes.

## Data Availability

All data produced in the present study are available upon reasonable request to the authors

## ETHICS STATEMENT

The study was conducted according to the principles of good clinical practice (GCP), along with the Declaration of Helsinki (1996), and in accordance with local regulations and the International Conference on Harmonization (ICH) guidelines. All study-related documents were approved by an independent institutional review board (IRB; MidLands Independent Review Board, Overland Park, Kansas). Written informed consent was obtained from all study subjects at screening prior to initiation of any study related procedures.

## ACKNOWLEDGEMENTS

We thank the study participants, the investigational team, laboratories, and reviewers; and our CRO partner Altasciences and their medical writing team.

## DISCLOSURES

Chris Galloway, MD, is the Chief Medical Officer at Diffusion Pharmaceuticals Inc.

## Notes

### Competing Interest Statement

The authors have declared no competing interest.

### Clinical Trial

NCT04808622

### Funding Statement

This study was funded by Diffusion Pharmaceuticals.

### Author Declarations

All study-related documents were approved by an independent institutional review board (IRB; Midlands Independent Review Board, Overland Park, Kansas)

